# Disrupted seasonality and association of COVID-19 with medically attended respiratory syncytial virus infections among young children in the US: January 2010–January 2023

**DOI:** 10.1101/2023.05.12.23289898

**Authors:** Lindsey Wang, Pamela B. Davis, Nathan A. Berger, David C. Kaelber, Nora D. Volkow, Rong Xu

## Abstract

Respiratory syncytial virus (RSV) infections and hospitalizations surged sharply in 2022 among young children. To assess whether COVID-19 contributed to this surge, we leveraged a real-time nation-wide US database of electronic health records (EHRs) using time series analysis from January 1, 2010 through January 31, 2023, and propensity-score matched cohort comparisons for children aged 0**–**5 years with or without prior COVID-19 infection. Seasonal patterns of medically attended RSV infections were significantly disrupted during the COVID-19 pandemic. The monthly incidence rate for first-time medically attended cases, most of which were severe RSV-associated diseases, reached a historical high rate of 2,182 cases per 1,0000,000 person-days in November 2022, corresponding to a related increase of 143% compared to expected peak rate (rate ratio: 2.43, 95% CI: 2.25**–**2.63). Among 228,940 children aged 0**–**5 years, the risk for first-time medically attended RSV during 10/2022**–**12/2022 was 6.40% for children with prior COVID-19 infection, higher than 4.30% for the matched children without COVID-19 (risk ratio or RR: 1.40, 95% CI: 1.27**–**1.55); and among 99,105 children aged 0**–**1 year, the overall risk was 7.90% for those with prior COVID-19 infection, higher than 5.64% for matched children without (RR: 1.40, 95% CI: 1.21**–**1.62). These data provide evidence that COVID-19 contributed to the 2022 surge of severe pediatric RSV cases.

## Introduction

Respiratory syncytial virus (RSV) is a leading cause of lower respiratory tract infection in young children^1^. The COVID-19 pandemic disrupted RSV and other respiratory viral infection patterns in the US for 2020**–**2021^2^. Unusually early and high rates of hospitalizations with RSV infections were reported in 2022, particularly among the youngest children^3,4^. However, the underlying reasons remain unknown. A recent study sequenced 105 RSV-positive specimens from symptomatic patients diagnosed with RSV during the autumn 2022 surge and showed that viral characteristics did not contribute to the extent or severity of the surge^5^, suggesting that non-viral influences on RSV transmission and severity may have contributed to that surge^3^. Nonpharmaceutical interventions such as masking and social distancing earlier in the pandemic prevented RSV from spreading and built a susceptible population with diminished immunity (“immunity debt”)^6–8^, which may have led to the large outbreaks in the 2021 winter in the US^9^ and in other countries^10^. However, the unusual surge of severe RSV cases in the winter of 2022 suggests that additional factors contributed. COVID-19 has long-lasting adverse effects on multiple organ systems, including immune, respiratory, endocrine, cardiovascular, neurological among others^11–15^. We hypothesize that COVID-19 contributed to the 2022 surge of severe pediatric RSV diseases, likely through its damage to the immune and respiratory systems of young children. Leveraging a nation-wide, real-time database of electronic health records (EHRs) of 61.4 million patients in the US, including 1.7 million children 0**–**5 years of age, we performed time series analyses to examine the time trend of medically attended RSV infections among young children in the US from January 1, 2010 through January 31, 2023. We then performed cohort studies to investigate whether prior COVID-19 infection was associated with increased risk for medically attended RSV infections while accounting for other risk factors.

## Results

### Seasonal pattern of monthly incidence rate of medically attended RSV infection among young children in the US during January 2010–January 2023

We examined EHR data from 29,013,937 medical visits for children aged 0**–**5 years and 13,169,955 medical visits from children aged 0**–**1 year from January 1, 2010 through January 31, 2023. From 2010 through 2019, the monthly incidence rate of medically attended RSV infection in children aged 0**–**5 years followed a consistent seasonal pattern: rising from September to November, peaking from December to January, then dropping from February to April, with sustained low rate during May to August (**Figure1a**).

**Figure 1.**
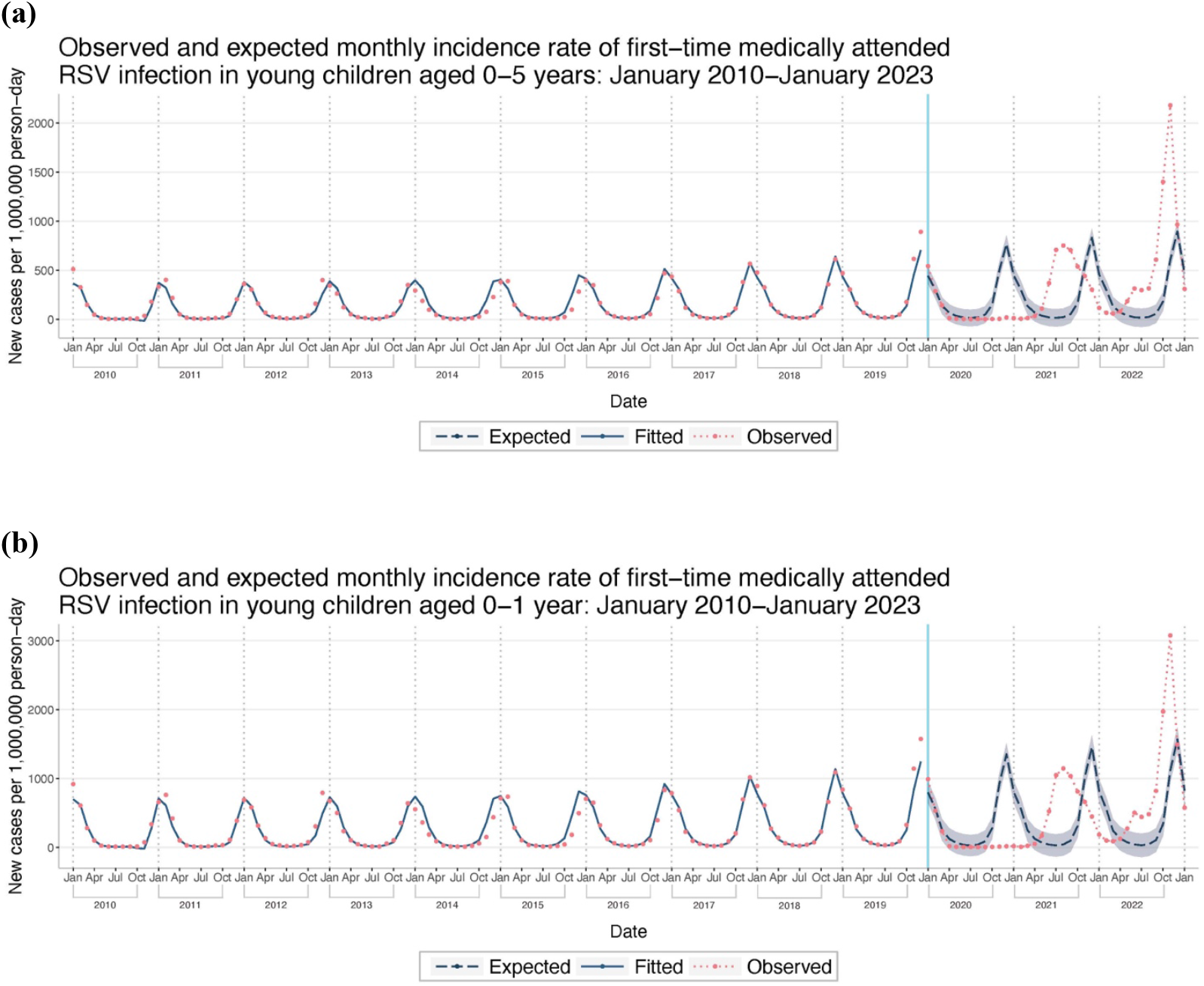
Observed and expected monthly incidence rate of first-time medically attended RSV infections from 1/1/2010 through 1/31/2023 among (a) children aged 0–5 years and (b) children aged 0–1 year. Monthly incidence rates were calculated as the number of incident cases per 1,000, 000 person-days for each month. The blue vertical line marks the beginning of COVID-19 pandemic (January 2020).

During the COVID-19 pandemic, the seasonal pattern changed. In 2020, the seasonal variation disappeared, and the incidence rate was consistently low from April through December ranging from 13 to 20 cases per 1,000,000 person-days. Compared with an expected December peak incidence rate of 770 cases per 1,000,000 person-days from the pre-pandemic trend, the incidence rate of 20 cases per 1,000,000 person-days in December 2020 corresponded with a relative decrease of 97.4% (Rate ratio or RR: 0.026, 95% CI: 0.017**–**0.040) (**Figure1a**).

In 2021, the seasonality returned but started earlier than expected. The incidence rate peaked in August, four months earlier than the expected peak in December. In August, the incidence rate was 753 cases per 1,000,000 person-days, corresponding with a relative decrease of 9.6% from the expected peak incidence rate (RR: 0.90, 95% CI: 0.82**–**0.99). The RSV season in 2021 extended to 9 months (May 2021**–**January 2022), longer than the expected 5 months in the winter (**Figure1a**).

In 2022 and January 2023, the seasonal pattern is similar to pre-pandemic years with a winter peak in November (2,182 cases per 1,000,000 person-days), one month earlier and 143% higher than the expected December peak (cases per 1,000,000 person-days) (RR: 2.43, 95% CI: 2.25**–** 2.63). The incidence rate in May through November 2022 was significantly higher than expected, corresponding to a related increase of 365%**–**1,576%. The incidence rate returned to the expected level in December (RR: 1.08, 95% CI: 0.98**–**1.18). In January 2023, the incidence rate was 311 cases per 1,000,000 person-days, 33% lower than the expected rate (RR: 0.67, 95% CI: 0.58**–**0.77) (**Figure1a**).

The incidence rate in children aged 0**–**1 year follow the same seasonal pattern as for children aged 0**–**5 years, but with higher rates. In 2022, the incidence rate peaked in November (3,077 cases per 1,000,000 person-days), representing a 96.4% increase from the expected December peak rate of 1,569 cases per 1,000,000 person-days (RR: 1.96, 95% CI: 1.85**–**2.09) (**Figure 1b**).

### Consistent time trend and seasonality between EHR-based incidence rate of medically attended RSV infection and CDC reported RSV-associated hospitalization among young children from October 2018–January 2023

The CDC RSV-NET is a population-based surveillance network for laboratory-confirmed RSV-associated hospitalizations in 12 states covering almost 8% of the US^16^, which began tracking RSV-associated hospitalizations in children during the 2018–2019 season. The time trend and seasonality of the EHR-based incidence rate of medically attended RSV infections in children aged 0–5 years (**Figure 2a**) is consistent with the CDC-reported RSV-associated hospitalization in children aged 0–4 years (**Figure 2b**): low RSV infection during 2020, early shift of the RSV season in 2021, surge in 2022 winter and decline in January 2023. In the pre-pandemic 2018**–** 2019 seasons, the peak of medically attended RSV infection preceded the RSV-associated hospitalizations. In the 2021–2022 seasons, the peaks were aligned.

**Figure. 2.**
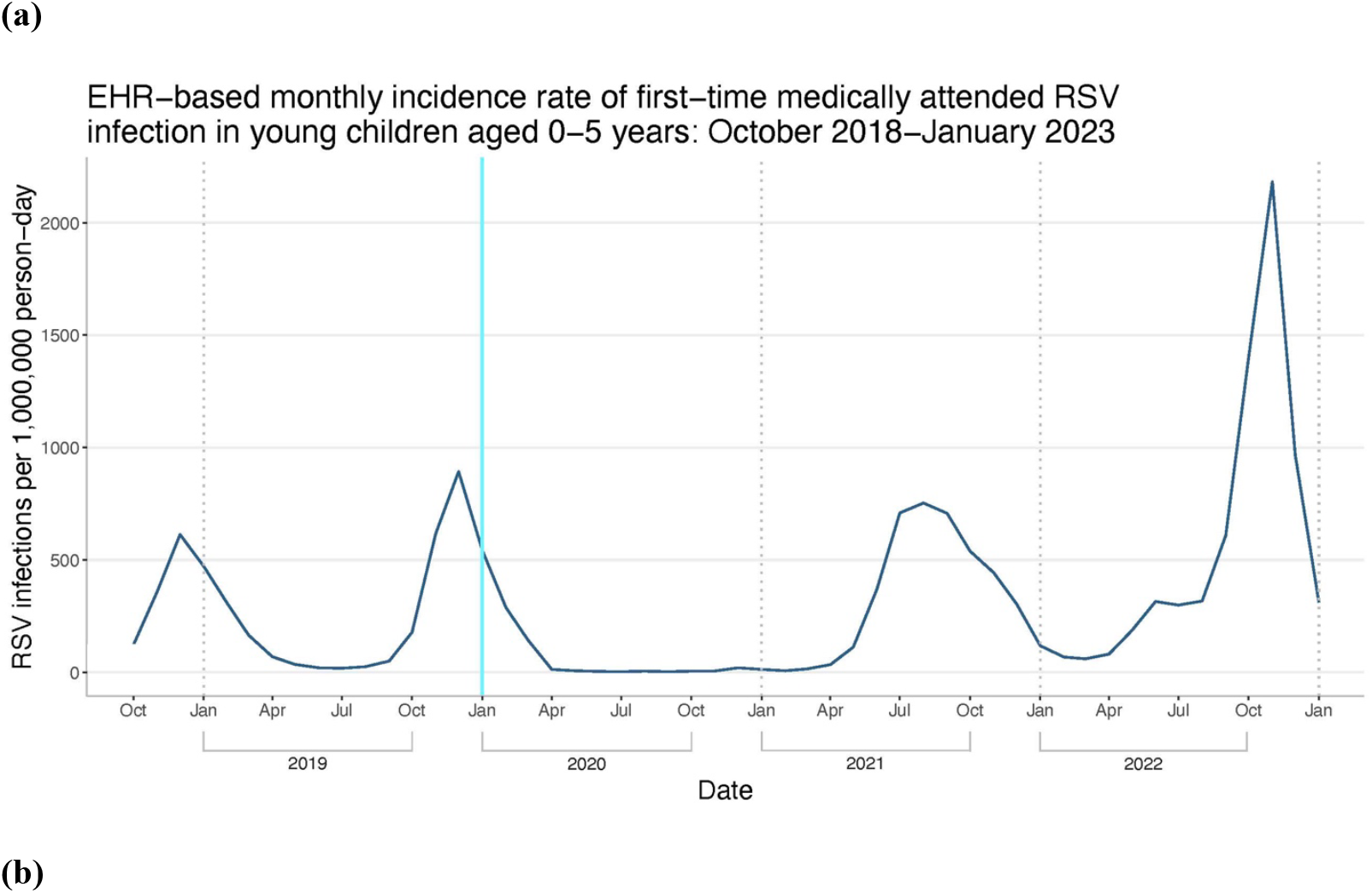
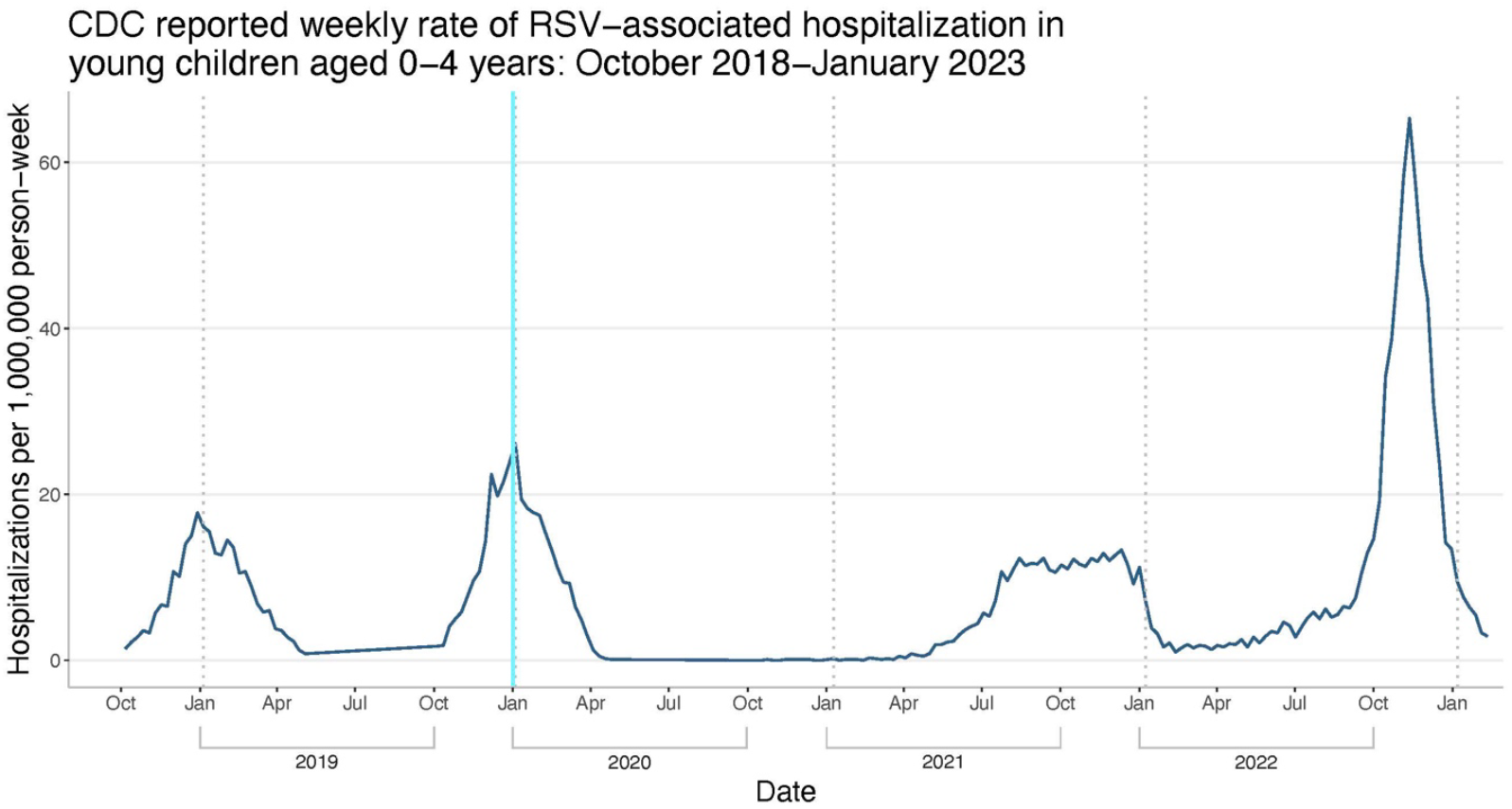
Time trend and seasonality of (a) EHR-based monthly incidence rate of first-time medically attended RSV infection among children aged 0–5 years from 10/1/2018–1/31/2023 and (b) the CDC reported rate of RSV-associated hospitalizations among young children aged 0– 4 years from 10/1/2018–1/31/2023.

### Stratified analysis by RSV severity: positive lab-test RSV infection, RSV-associated diseases, RSV-associated bronchiolitis, and unspecified bronchiolitis

In this study, medically attended RSV infection was defined by 12 laboratory test codes and 3 disease clinical diagnosis codes. The clinical diagnosis-based RSV infection included “Respiratory syncytial virus pneumonia” (J12.1), “Acute bronchiolitis due to respiratory syncytial virus” (J21.0), and “Respiratory syncytial virus as the cause of diseases classified elsewhere” (B97.4). We performed stratified analyses for 3 types of RSV cases based on: clinical diagnosis (J12.1, J21.0, B97.4), positive lab tests (12 lab test codes) and RSV-associated bronchiolitis (J21.0). In addition, we also examined unspecified acute bronchiolitis (J21.9), which could be caused by other severe respiratory viruses such as influenza.

During the 2022 peak season in November and December, clinically diagnosed RSV had the highest incidence rate in children 0**–**5 years old, followed by positive lab test-confirmed RSV and RSV-associated bronchiolitis, all reaching a historically high rate. For comparison, the peak incidence rate of unspecified bronchiolitis in 2022 was similar to that during the pre-pandemic period of 2015**–**2019 but higher than in 2020 and 2021 (**Figure 3a**). Starting in 2015, there was a marked increase in incidence rate of unspecified bronchiolitis cases (**Figure 3a**). Among children 0**–**1 year old, clinically diagnosed RSV diseases had the highest incidence rate during the 2022 RSV peak season, followed by RSV-associated bronchiolitis and positive lab test-confirmed RSV, all of which reached a historically high rate (**Figure 3b**). The rate of unspecified bronchiolitis in 2022 was similar to that during the pre-pandemic period of 2015**–**2019. In summary, these stratified analyses of different indicators showed differential trends between RSV infection and non-RSV respiratory viral diseases, indicating the specificity of the 2022 surge in RSV infection children.

**Figure 3.**
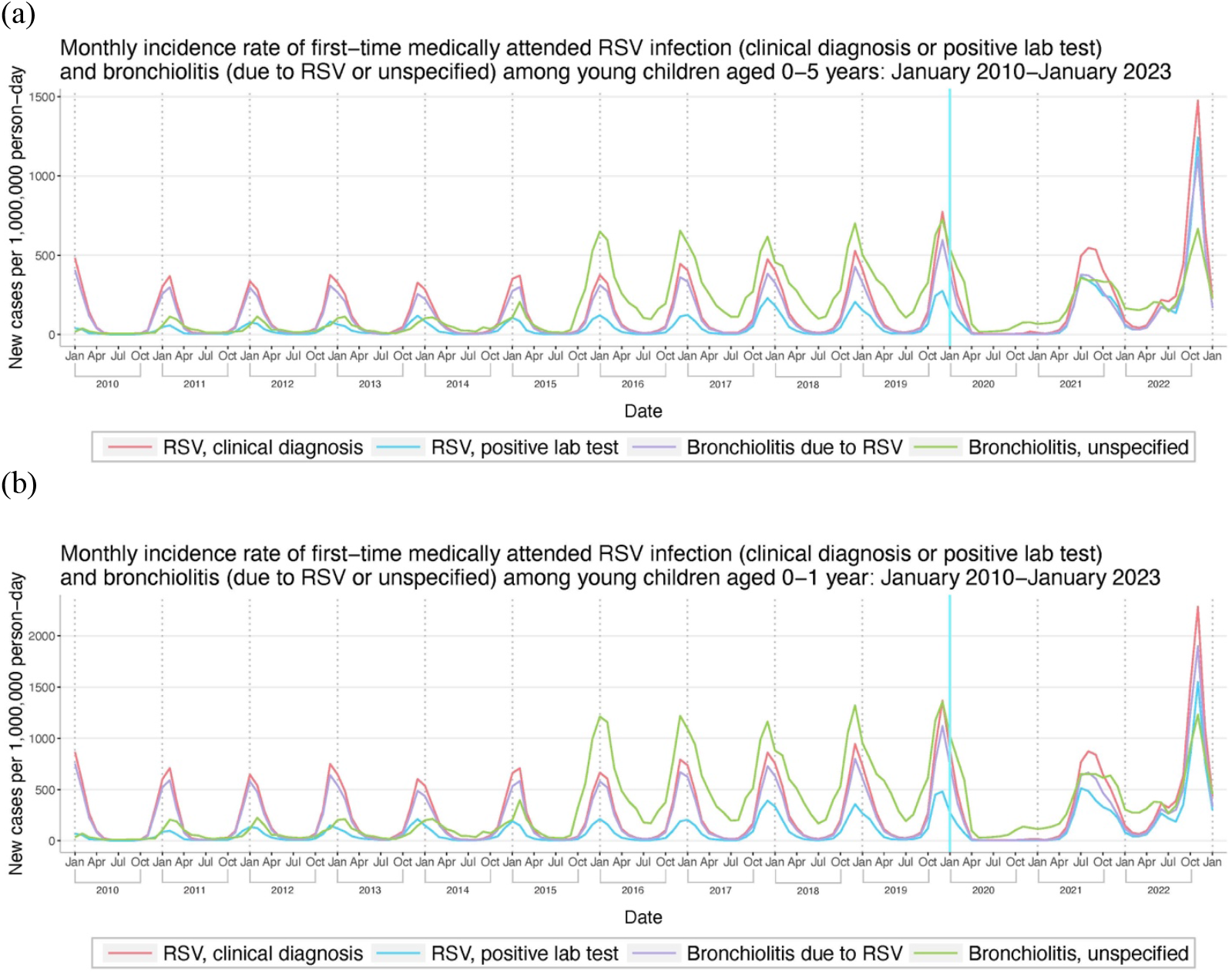
Stratified analysis of monthly incidence rate of first-time medically attended RSV infection from 1/1/2010 through 1/31/2023 among (a) children aged 0–5 years and (b) children aged 0–1 year. Monthly incidence rates were calculated as the number of incident cases per 1,000,000 person-days for each month. The blue vertical line marks the beginning of COVID-19 pandemic (January 2020). RSV infection was stratified by clinical diagnosis (J12.1, J21.0, B97.4), positive lab tests (12 lab-test codes), RSV-associated bronchiolitis (J21.0). Unspecified acute bronchiolitis (J21.9), which could be caused by other severe respiratory viruses such as influenza, was also shown.

### COVID-19 is associated with a significant increased risk for first-time medically attended RSV infection among young children during peak seasons in 2022 and 2021

We then examined if prior COVID-19 infection was associated with increased risk of new diagnosis of medically attended RSV infection among young children during both the 2022 and 2021 peak seasons. To examine the association of prior COVID-19 infection and first-time RSV infection in 2022 peak season (October–December) among young children, the study population comprised 228,940 children aged 0–5 years (age as of October 2022) who had medical encounters with healthcare organizations in 10/2022 and had no prior medically attended RSV infection. The study population included 14,493 children who contracted COVID-19 prior to 8/2022 (“COVID-19 (+) cohort”) and 214,447 children who had no EHR-documented COVID-19 infection (“COVID-19 (–) cohort”). Compared to the COVID-19 (–) cohort, the COVID-19 (+) cohort was older and had significantly higher prevalence of adverse socioeconomic determinants of health, pre-existing medical conditions, procedures and COVID-19 vaccination (**Table 1**). After propensity-score matching, the two cohorts (14,488 children in each) were balanced (**Table 1**).

**Table 1:** Characteristics of the 2022 study cohorts of children aged 0–5 years (as of 10/2022) who had a medical encounter with healthcare organizations in 10/2022 and had no prior medically attended RSV infection before and after propensity-score matching for the listed variables.

|  | **Before Propensity-Score Matching** |  |  | **After Propensity-Score Matching** |  |  |
| --- | --- | --- | --- | --- | --- | --- |
|  | **COVID-19 (+) cohort** | **COVID-19 (–) cohort** | **SMD** | **COVID-19 (+) cohort** | **COVID-19 (–) cohort** | **SMD** |
| **Total number** | 14,493 | 214,447 |  | 14,488 | 14,488 |  |
| **Age at index event (years, mean±SD)** | 2.4± 1.6 | 2.1± 1.8 | 0.15* | 2.4± 1.6 | 2.4± 1.6 | 0.008 |
| **Sex (%)** |  |  |  |  |  |  |
| Female | 46.8 | 46.9 | 0.003 | 46.8 | 46.8 | 0.001 |
| Male | 53.2 | 53.1 | 0.003 | 53.2 | 53.2 | 0.001 |
| **Ethnicity (%)** |  |  |  |  |  |  |
| Hispanic/Latinx | 22.0 | 20.6 | 0.03 | 22.0 | 21.8 | 0.005 |
| Not Hispanic/Latinx | 62.5 | 59.5 | 0.06 | 62.4 | 62.7 | 0.005 |
| Unknown | 15.6 | 19.9 | 0.11* | 15.6 | 15.5 | 0.001 |
| **Race (%)** |  |  |  |  |  |  |
| Asian | 3.2 | 3.5 | 0.01 | 3.2 | 3.1 | 0.005 |
| Black | 15.5 | 15.6 | 0.002 | 15.5 | 15.5 | <.001 |
| White | 60.7 | 57.9 | 0.06 | 60.7 | 61.2 | 0.01 |
| Unknown | 20.1 | 22.4 | 0.06 | 20.1 | 19.7 | 0.01 |
| **Adverse socioeconomic determinants of health (%)** | 7.5 | 3.8 | 0.16* | 7.4 | 7.6 | 0.006 |
| **Pre-existing medical conditions and treatments (%)** |  |  |  |  |  |  |
| Diseases related to blood and immune mechanisms | 13.6 | 6.4 | 0.24* | 13.5 | 13.7 | 0.005 |
| Diseases related to immune mechanisms | 2.3 | 0.6 | 0.14* | 2.3 | 2.2 | 0.007 |
| Chronic lower respiratory diseases | 8.6 | 4.1 | 0.19* | 8.5 | 8.6 | 0.002 |
| Chronic lower respiratory diseases originating in the prenatal period | 1.4 | 0.8 | 0.06 | 1.4 | 1.2 | 0.02 |
| Asthma | 7.6 | 3.7 | 0.17* | 7.6 | 7.7 | 0.004 |
| Down syndrome | 0.6 | 0.4 | 0.04 | 0.6 | 0.4 | 0.02 |
| Malnutrition | 2.0 | 0.8 | 0.11* | 1.9 | 1.6 | 0.03 |
| Disorders of newborn related to length of gestation and fetal growth | 8.4 | 4.7 | 0.15* | 8.4 | 8.5 | 0.005 |
| Neoplasms | 7.5 | 3.8 | 0.16* | 7.5 | 7.4 | 0.002 |
| Congenital malformations of the circulatory system | 6.4 | 4.0 | 0.11* | 6.4 | 6.1 | 0.01 |
| Heart diseases | 5.1 | 2.3 | 0.15* | 5.1 | 4.9 | 0.01 |
| Neuromuscular Disorders | 0.2 | 0.0 | 0.04 | 0.2 | 0.2 | 0.02 |
| Chemotherapy | 2.1 | 0.8 | 0.10* | 2.1 | 1.9 | 0.02 |
| COVID-19 vaccines | 9.3 | 4.6 | 0.21* | 9.3 | 9.3 | 0.002 |

COVID-19 (+) cohort – children who contracted COVID-19 prior to 8/2022 as documented in their EHRs. COVID-19 (–) cohort – children who had no documented COVID-19 in their EHRs. Index event was a medical visit in 10/2022. Cohorts were propensity-score matched for the list variables with variable status based on anytime to 1 day before the index event. The status of adverse socioeconomic determinants of health (SDOHs) was based on the ICD-10 code “Persons with potential health hazards related to socioeconomic and psychosocial circumstances” (Z55-Z65), which includes codes “Problems related to housing and economic circumstances” (Z59), “Problems related to upbringing” (Z62), among others. SMD – standardized mean differences. \*SMD greater than 0.1, a threshold recommended for declaring imbalance.

Both cohorts were followed for 60 days starting from a medical visit in 10/2022. The overall risk for first-time medically attended RSV infection during 10/2022–12/2022 was 6.04% for the COVID-19 (+) cohort, higher than the 4.30% for the propensity-score matched COVID-19 (–) cohort (risk ratio or RR: 1.40, 95% CI: 1.27-1.55), with highest association for clinically diagnosed RSV diseases (RR: 1.44, 95% CI: 1.27–1.63) including RSV-associated bronchiolitis (RR: 1.43, 95% CI: 1.23–1.67) (**Figure 4**). Prior COVID-19 infection was associated with a significantly increased risk for unspecified bronchiolitis (RR: 1.26, 95% CI: 1.05–1.53).

**Figure 4.**
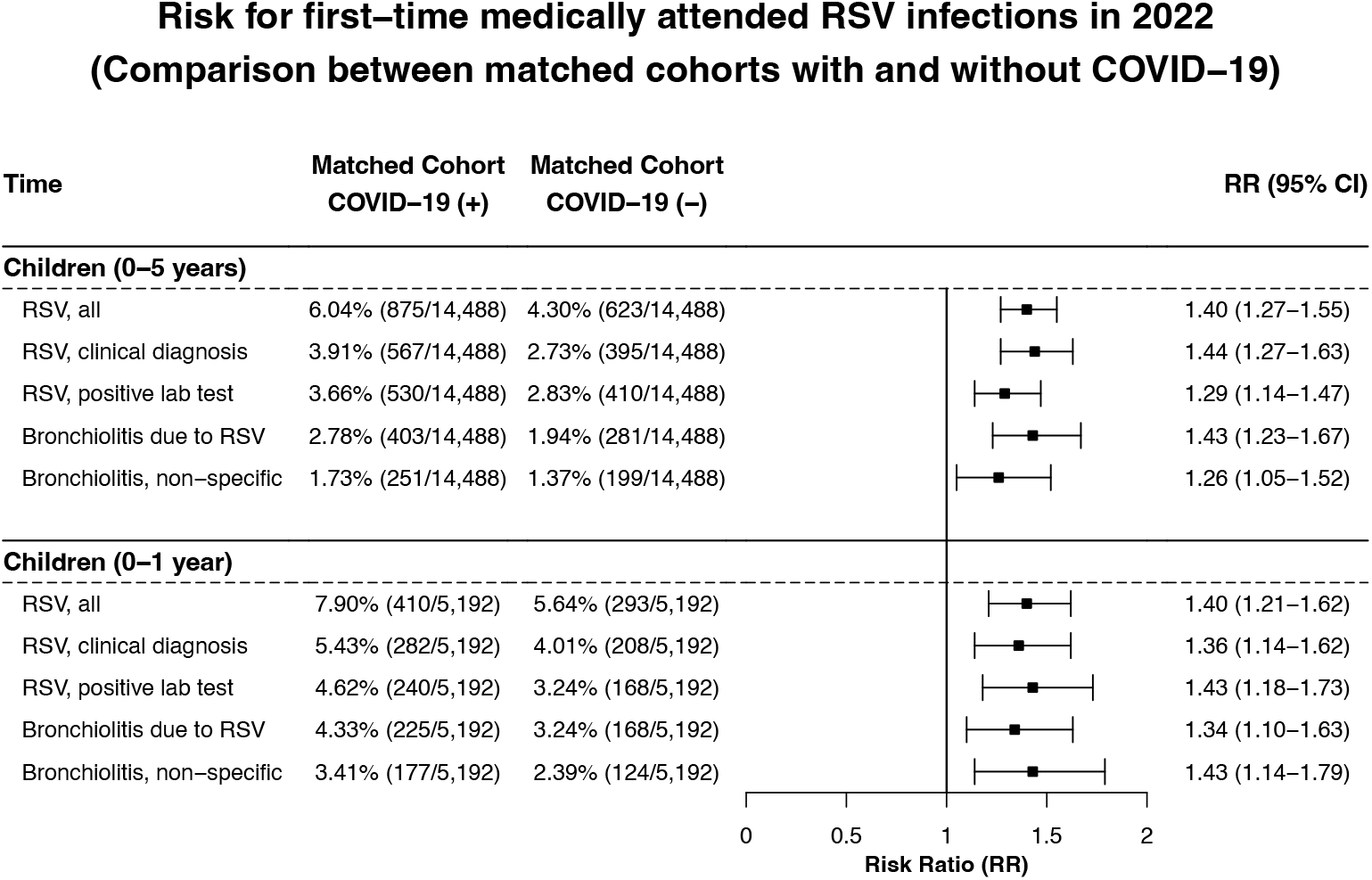
Comparison of risk for first-time medically attended RSV infection that occurred during the 2022 RSV peak season (October–December) among young children who had medical encounters with healthcare organizations in 10/2022 and had no prior medically attended RSV infection. COVID-19 (+) cohort – children who contracted COVID-19 prior to 8/2022 as documented in their EHRs. COVID-19 (–) cohort – children who had no documented COVID-19 in their EHRs. Index event was a medical visit in 10/2022. Outcomes (RSV infections) were followed 0-60 days starting from the index event and occurred during 10/1/2022–12/31/2022. Cohorts were propensity-score matched for demographics (actual age at index event, gender, race, ethnicity), adverse socioeconomic determinants of health, pre-existing medical conditions and procedures and COVID-19 vaccination.

Among the population of 228,940 children aged 0–5 years, 99,105 children were aged 0–1year (as of October 2022), including 5,193 with prior COVID-19 infection (COVID (+) cohort) and 93,912 without (COVID-19 (–) cohort). Compared to the COVID-19 (–) cohort, the COVID-19 (+) cohort was older and had higher prevalence of adverse socioeconomic determinants of health, pre-existing medical conditions, and COVID-19 vaccinations. After propensity-score matching, both cohorts comprised 5,192 children and were balanced (data not shown). The overall risk for RSV among children aged 0–1 year during 10/2022–12/2022 was 7.90% for the COVID-19 (+) cohort, higher than 5.64% for the matched COVID-19 (–) cohort (RR: 1.40, 95% CI: 1.21–1.62). Increased risks were observed for severe RSV diseases, positive lab test-confirmed RSV infection, and unspecified bronchiolitis (**Figure 4**).

In 2021, the masking and social distancing policy was still in place to maximize protection from the Delta variant, although lockdowns eased. However, unlike 2020, the observed incidence rate of RSV infection in 2021 reached pre-pandemic level and the season extended to 9 months (**Figure 1**), suggesting that young children in 2021 were more vulnerable to RSV infections and more children contracted RSV than the pre-pandemic population. The 2021 study population comprised 370,919 children aged 0–5 years and 162,771 children aged 0–1 year (age as of 7/2021–8/2021. By comparing propensity-score matched COVID-19 (+) and COVID-19 (-) cohorts, we showed that prior COVID-19 infection was associated with increased risk for first-time medically attended RSV infection during the 2021 RSV season (7/2021–12/2021) among children aged 0–5 years (RR: 1.32, 95% CI: 1.12–1.56) and children 0–1 year (RR: 1.47, 95% CI: 1.18–1.82) (**Figure 5**). COVID-19 was associated with a significantly increased risk for severe RSV diseases, positive lab test-confirmed RSV and unspecified bronchiolitis in 2021.

**Figure 5.**
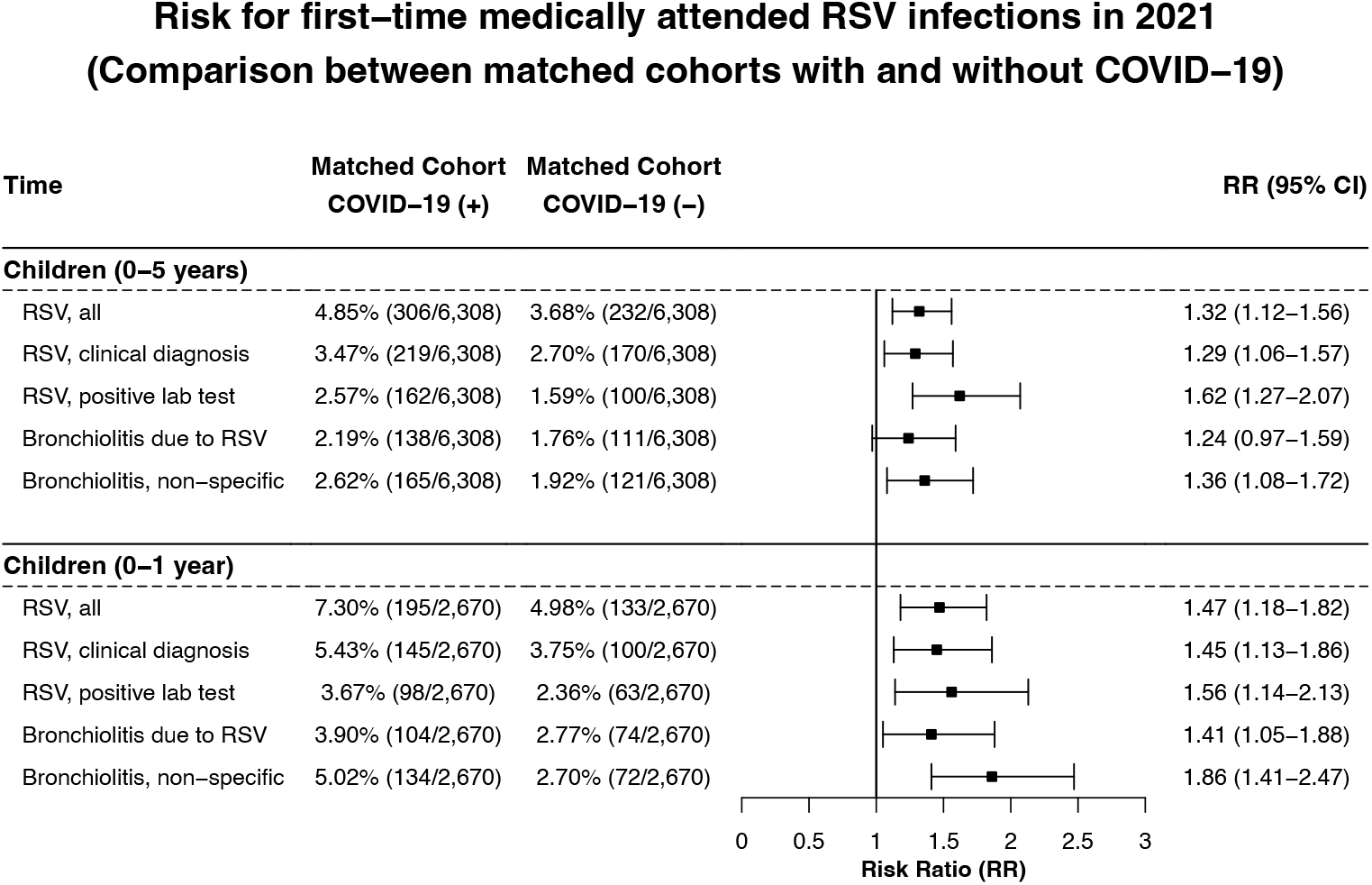
Comparison of risk for medically attended RSV infection that occurred during the 2021 RSV peak season (July–December) among young children who had medical encounters with healthcare organizations from 7/2021–8/2021and had no prior medically attended RSV infection. COVID-19 (+) cohort – children who contracted COVID-19 prior to 6/1/2022 as documented in their EHRs. COVID-19 (–) cohort – children who had no documented COVID-19 in their EHRs. Index event was a medical encounter during 7/2021-8/2021. Outcomes (RSV infections) were followed 0-120 days starting with the index event and occurred from 7/2021 through 12/2021. COVID-19 (+) and COVID-19 (–) cohorts were propensity-score matched for demographics (actual age at index event, gender, race, ethnicity), adverse socioeconomic determinants of health, pre-existing medical conditions, and procedures.

## Discussion

In this study, we focused on medically attended RSV infections, which was based on the presence of clinical diagnosis codes for RSV-associated diseases including pneumonia and bronchiolitis or positive lab test results for RSV infections in a patient’s EHR that required medical care or hospitalization. Children with mild infections are often not sick enough to present to a healthcare setting. Even in a healthcare setting, most providers do not test for RSV unless the patient is being hospitalized. Among the 19,936 incidence cases of RSV infections occurred in children aged 0–5 years during 10/2022-12/2022, 14,103 (70.7%) were severe RSV-associated diseases including RSV-associated bronchiolitis (54.5%). Based on EHR data alone, it is difficult to discern whether two close RSV visits recorded in patient EHRs were for two different or the same RSV infection. In this study, we used first-time medically attended RSV infections in order to not double count two different clinical visits for the same RSV infection.

In 2020, the incidence rate of RSV infection was low throughout the year, which is likely due to nonpharmaceutical interventions such as lockdown, masking and social distancing that prevented RSV from spreading. Computational models that simulated nonpharmaceutical interventions and associated immunity debt predicted a large outbreak in the 2021 winter^6,9^. However, our data showed that the seasonal pattern of medically attended RSV infection in children aged 0–5 years returned but the incidence rate was lower for severe diseases including RSV-associated bronchiolitis than expected. However, the RSV season in 2021 extended to 9 months, resulting in more RSV-infected children in 2021 than in pre-pandemic. Positive lab test-confirmed RSV infection reached pre-pandemic levels for children aged 0–1 year and was higher for children aged 0–5 years. Although there was a buildup of susceptible children in 2021, certain COVID-19 preventative measures remained in place in 2021 that limited the spread of RSV infections. In April 2022, the CDC lifted the mask mandate but still recommended that people wear masks at public transportation settings^17^. Interestingly, very young children aged 0-1 year (as of 2021), many of whom were born after 2020, also showed increased RSV infection in 2021. Waning maternal immunity due to low RSV exposure during the COVID-19 pandemic and the consequent decrease in transplacental RSV antibody transfer may have contributed to increased RSV infections in 2021 compared to 2020. Our cohort studies comparing the matched COVID-19 (+) and COVID-19 (–) cohorts for children aged 0–5 years and children aged 0–1 year showed that prior COVID-19 infection was associated with an increased risk for RSV infection including both clinically diagnosed RSV diseases and positive lab test-confirmed RSV infection in 2021. However, RSV infections in 2021 did not reach the levels in 2022, largely because of the preventive measures and fewer COVID-19-infected children.

In 2022, medically attended RSV infection including both clinically diagnosed severe RSV diseases and positive lab test-confirmed infections reached a historically high rate, higher for severe RSV diseases than the positive lab test-confirmed RSV infection. Among children aged 0–1, the peak incidence rate of severe RSV-associated diseases in November 2022 was 2,285 cases per 1,000,000 person-days, 47% higher than lab test-confirmed RSV. These data suggest that the 2022 RSV surge was disproportionately driven by more severe cases of RSV diseases, which could not be fully explained by increased testing practices, awareness, or transmission through day-care or siblings alone. While nonpharmaceutical interventions in 2021 and immunity debt contributed to the increased rate of RSV infection, these factors alone could not fully explain the huge surge in November 2022, representing 4–5 times as many severe RSV-associated diseases as in 2021. For children aged 0–1 year (as of 2022), if the immune debt due to waning maternal immunity was the main contributor, we would expect that the level of RSV infection in 2022 to similar to that in 2021. Instead, the peak incidence rate of severe RSV diseases in children aged 0-1 year was 2,285 cases per 1,000,000 person-days in November 2022, a 161% increase compared to the peak rate of 874 cases per 1,000,000 person-days in August 2021. The comparisons between matched COVID-19 (+) and COVID-19 (–) cohorts showed that prior COVID-19 infection was associated with increased risk for RSV infection in 2022. In 2022, significantly more children contracted COVID-19^18^ due to the relaxation of preventive measures and the dominance of the highly transmissible Omicron variant^19^. Together with the effects of RSV-specific immunity debt and other factors, the large buildup of COVID-19-infected children and the potential long-term adverse effects of COVID-19 on the immune and respiratory systems^11–15^ may have contributed to the 2022 winter surge of severe RSV diseases.

RSV is the most common virus associated with bronchiolitis. However, many other viruses cause bronchiolitis, including human rhinovirus, coronavirus, metapneumovirus and adenovirus^20^. In the winter of 2015, there was a marked increase in incidence rate of unspecified bronchiolitis, which remained stable from 2015 through 2019. Starting in 2017–2018, there was an increase in the incidence rate of lab test-confirmed RSV but was not accompanied by a proportional increase in clinically diagnosed RSV diseases until in 2019–2020. The underlying reasons for these changes remain unknown, but may be attributable to advances in genomic surveillance, increased detection of respiratory viruses in the laboratory, the emergence of new viral strains, increased awareness, among others. The linear time series regression model used a linear term to model the increasing time trend, however the observed peak incidence rate of RSV in 2022 was 143% higher than expected, suggesting that these factors may not be the sole contributing factor for the 2022 surge.

We show that the incidence rate of unspecified bronchiolitis surged in November 2022, higher than in 2020 and 2021, but was similar or lower than in 2018–2019. The cohort studies showed that prior COVID-19 infection was associated with increased risk for unspecified bronchiolitis in both 2021 and 2022. Patients infected with COVID-19 can have long-lasting changes in both innate and lymphocyte based immune functions^11,12,17-19^, precisely the systems most engaged in defense against respiratory viruses^21,22^. These findings further support our hypothesis that COVID-19 had adverse impact on the immune and respiratory systems of children, making them susceptible to severe respiratory viral infections from RSV and other viruses. While COVID-19 infection was associated with an increased risk for unspecified bronchiolitis, we did not observe a historically high surge in 2022, likely because unspecified bronchiolitis was not as common as RSV infection. Findings from our study could offer a unique opportunity to further understand the mechanisms of SARS-CoV-2 viral infection, RSV infection, other respiratory viruses, and their potential positive interactions^23^.

Several studies from the US and other countries reported disrupted seasonality of RSV infection during 2020–2021^2,10,24,25^. Our study is the first to report the US nation-wide time series data of first-time medically attended RSV infection among young children from January 2010 through January 2023. The CDC RSV-NET is a population-based surveillance network for laboratory-confirmed RSV-associated hospitalizations in 12 states^16^, which began tracking in children in 2018–2019. We show that our EHR-based incidence rate of f medically attended RSV infection corresponds closely with the CDC reported RSV-associated hospitalizations. In addition, our study provides information that is complementary to the CDC data. Firstly, the EHR-based data reported medically attended RSV cases, the majority of which were severe RSV-associated diseases including bronchiolitis. On the other hand, the CDC reported hospitalizations associated with laboratory-confirmed RSV. Secondly, our study population was drawn from a nation-wide database of EHRs collected in diverse clinical settings (inpatient, outpatient, emergency) across all 50 states in the US, representing demographically and clinically diverse RSV cases. The database also contains longitudinal patient data of more than 20 years, which allowed us to build robust time-series models. On the other hand, the CDC RSV-NET began data collection in children in 2018 based on voluntary reporting from state health departments. The EHR database is updated daily, and RSV-NET is updated weekly. Thirdly, the EHR data contain rich information of patients including demographics, medical conditions, procedures, medications, lab tests and socio-economic determinants of health. These additional data allowed us to further examine if prior COVID-19 infection was associated with an increased risk for RSV infection that accounted for the RSV surge in 2022 while accounting for other risk factors. Although each data resource has inherent limitations, their consistent trends reinforce the validity of our findings. Together, these complementary data resources provide a comprehensive overview of updated RSV activity and outcomes at both medical and general population levels in the US.

The findings comparing the propensity matched cohorts showed that prior COVID-19 infection was associated with a significant increased risk for RSV infection during both 2022 and 2021 RSV peak seasons. This finding is consistent with our hypothesis that COVID-19 is an important contributing factor for the 2022 surge of severe pediatric RSV diseases, likely through its lasting damage to the immune and respiratory systems of young children. Although the strength of the associations in 2022 are similar to that in 2021, we observed historically high monthly incidence rates of severe RSV cases only in 2022, but not in 2021. However, the RSV season in 2021 was extended to 9 months. Nearly as many children were infected with RSV in 2021 as in 2022 (25,638 and 29,388 incidence cases in May–December 2021 and 2022, respectively). Although it is plausible that similar factors placed children at risk for both medically attended COVID-19 and RSV, our study controlled for as many known risk factors as possible. To further mitigate potential confounding effects, we put an extra restriction on the relative timing of prior COVID-19 infection and RSV infection for the COVID-19 (+) cohort: COVID-19 occurred at least 2 months prior to RSV infection. However, this is an associational study that could suffer from other uncontrolled or unmeasured confounders. Further mechanistic studies are needed to investigate how COVID-19 promotes severe RSV diseases in young children.

The finding that COVID-19 is associated with increased risk for unspecified bronchiolitis further supports our hypothesis that COVID-19 renders children more susceptible to severe respiratory viral infections. Since COVID-19 has long-term effects on multiple organ systems in diverse populations we expect that it will also be associated with increased risk for other severe respiratory viral infections or other bacterial or viral infections in other populations including adults, older adults, immunocompromised patients (i.e., cancer, HIV, receiving immunosuppressive treatments) and people with underlying medical conditions.

Our study has several limitations: First, it focused on medically attended RSV infection. Although we stratified RSV infection based on positive lab-test and clinical diagnoses, due to restrictions from TriNetX we were unable to assess RSV and its outcomes (e.g., hospitalization) in different clinical settings (e.g., inpatient, outpatient, emergency). Second, the patients from the TriNetX network are those who had medical encounters with healthcare systems contributing to TriNetX. Therefore, they do not necessarily represent the entire US population. Although our study showed a time trend that is well-aligned with that from the CDC population-based surveillance data, results from this study need to be validated in other populations. Third, the EHR data were drawn from 34 healthcare organizations across all 50 states, covering diverse geographic regions. However, we were unable to further break down the trend patterns by region due to TriNetX’s de-identification restrictions. Fourth, many children have contracted COVID-19 though the actual prevalence is unknown^18^. The status of prior COVID-19 in our study was based on the clinical diagnosis code or positive lab test results captured in EHRs, which very likely was an underestimate of the actual rate because many COVID-19 tests were performed at home. This means that the COVID-19 (–) cohorts in our study might have included children with mild COVID-19 that were not documented in their EHRs. This could have underestimated the associations of COVID-19 with RSV infection reported in our study. Fifth, there may be over-/mis-/under-diagnosis of RSV infection and other diseases in patient EHRs. However, we compared the relative risk for RSV infection between cohorts drawn from the same TriNetX dataset, therefore these issues should not substantially affect the comparative risk analyses. Sixth, the COVID-19 (+) and COVID-19 (–) cohorts were matched for age, gender, ethnicity, race, adverse socio-economic determinants of health (including physical, social and psychosocial environment, and housing), pre-existing medical conditions, procedures and COVID-19 vaccinations. Among risk factors for RSV infection among young children^26^, day-care attendance and presence of older siblings in school or day-care may also be risk factors for SARS-CoV-2 viral transmission^27^. While patient EHRs did not capture such information and these uncaptured risk factors could represent unmeasured confounders, they alone could not explain the 2022 surge that was disproportionately driven by more severe cases of RSV diseases. Future studies are needed to examine the associations of COVID-19 and RSV in adults, which are not as often confounded by factors such as presence of siblings or schooling. In addition, although we have controlled for COVID-19 vaccination between cohorts, we were unable to assess how vaccination further modified the associations of COVID-19 with RSV due to small sample sizes as only 4.9% of our 2022 study population were vaccinated.

In conclusion, our study established the utility of EHRs as a cost-effective alternative for real-time pathogen and syndromic surveillance of unexpected disease patterns including RSV infection. Findings from our study support that prior COVID-19 infection was associated with a significantly increased risk for RSV infection and was a driving force for the 2022 surge of severe pediatric RSV cases in the US.

## Data Availability

This study used population-level aggregate and de-identified data generated by the TriNetX Platform. Due to data privacy, patient-level data were not used and cannot be shared. All the results related to this study are included in the manuscript.

## Acknowledgments

We acknowledge support from the National Institute on Aging (grants nos. AG057557, AG061388, AG062272, AG07664), National Institute on Alcohol Abuse and Alcoholism (grant no. AA029831), the Clinical and Translational Science Collaborative (CTSC) of Cleveland (grant no. TR002548-01), National Cancer Institute Case Comprehensive Cancer Center (CA221718, CA043703).

## Author information

### Contributions

R.X. conceptualized the project, designed and supervised the study. L.W. performed data analysis and prepared tables and figures. R.X. and L.W. drafted manuscript. P.B.D., N.A.B., D.C.K., N.D.V. critically contributed to result interpretation and manuscript preparation. We confirm the originality of content. R.X. had full access to all the data in the study and takes responsibility for the integrity of the data and the accuracy of the data analysis.

## Methods

### 1. Data

The data used in this study were collected during February 22**–**March 5, 2023 from the TriNetX “Research USA No Date Shift” Network. We used the TriNetX platform to access aggregated and de-identified electronic health records (EHRs) of 61.4 million patients including 1.7 million children 0–5 years of age from 34 health care organizations in the US across all 50 states, covering diverse geographic regions (31% in the Northeast, 13% in the Midwest, 42% in the South, and 13% in the West), age, race/ethnic, income and insurance groups and clinical setting (inpatient, outpatient, emergency, virtual)^1^. TriNetX, LLC is compliant with the Health Insurance Portability and Accountability Act (HIPAA). Any data displayed on the TriNetX Platform in aggregate form, or any patient level data provided in a data set generated by the TriNetX Platform only contains de-identified data as per the de-identification standard defined in Section §164.514(a) of the HIPAA Privacy Rule. TriNetX built-in analytic functions (e.g., incidence, prevalence, outcomes analysis, survival analysis, propensity score matching) allow for patient-level analyses, while only reporting population level data. The MetroHealth System, Cleveland OH, IRB determined research using TriNetX, in the way described here, is not Human Subject Research and therefore IRB exempt. We previously used the TriNetX platform to perform retrospective cohort studies and incidence rate analyses related to COVID-19 in various populations^2–13^ including in young children^2,10^.

TriNetX is a platform that de-identifies and aggregates electronic health record (EHR) data from contributing healthcare systems, most of which are large academic medical institutions with both inpatient and outpatient facilities at multiple locations, across all 50 states in the US. TriNetX Analytics provides web-based and secure access to patient EHR data from hospitals, primary care, and specialty treatment providers, covering diverse geographic locations, age groups, racial and ethnic groups, income levels and insurance types including various commercial insurances, governmental insurance (Medicare and Medicaid), self-pay/uninsured, worker compensation insurance, military/VA insurance among others.

Self-reported sex (female, male), race and ethnicity data in TriNetX comes from the underlying clinical EHR systems of the contributing healthcare systems. TriNetX maps race and ethnicity data from the contributing healthcare systems to the following categories: (1) Race: Asian, American Indian or Alaskan Native, Black or African American, Native Hawaiian or Other, White, Unknown race; and (2) Ethnicity: Hispanic or Latino, Not Hispanic or Latino, Unknown Ethnicity.

TriNetX completes an intensive data preprocessing stage to minimize missing values. All covariates are either binary, categorical (which expands to a set of binary columns), or continuous but essentially guaranteed to exist. Age is guaranteed to exist. Missing sex values are represented using “Unknown Sex”. The missing data for race and ethnicity are presented as “Unknown race” or “Unknown Ethnicity”. For other variables including medical conditions, procedures, lab tests and socio-economic determinant health, the value is either present or absent so “missing” is not pertinent.

### 2. Statistical analysis of seasonality

#### Incidence rate analysis

Monthly incidence rate of first-time medically attended RSV infection from January 1, 2010 through January 31, 2023 were examined in: (1) children aged 0**–**5 years who had any medical visits (including inpatient, outpatient, emergency) within the TriNetx network healthcare organizations and (2) children 0–1 year who had medical visits (npatient, outpatient, emergency) within the TriNetX Network of healthcare organizations. Age was determined as of the calendar month of the medical visit. For example, there were 224,702 medical visits and 1,256 RSV incidence cases in November 2016 in children aged 0**–**5 years (age as of November 2016). In November 2022, there were 266,086 medical visits and 9,923 RSV incidence cases by children aged 0**–**5 years (age as of November 2022). The status of RSV infection was based on 12 lab test codes and 3 disease clinical diagnosis codes.

We used the TriNetX built-in functions “Incidence & Prevalence analytic” to calculate incidence rate. For a given time window (each calendar month in our study), the incidence proportion denominator includes all and only those patients in the cohort under analysis (children ≤ 5 years of age as of that month when they had a medical visit), whose fact record overlaps the time window by at least one day, whose fact record does not contain the event of interest (RSV in this study) during the lookback period. For this study, the lookback time was any time to one day before the start of each month, therefore we examined first-time medically attended RSV infections. The incidence proportion numerator includes all and only those patients who are in the denominator and whose record includes the event of interest on a date within the time window. For each month, the incidence rate denominator is the product of the number of patients in the incidence proportion denominator and the number of days covered by the time interval. The incidence rate numerator is equivalent to the incidence proportion numerator.

#### Time series analysis

We used a linear time series regression model to model the pre-pandemic (January 1, 2010 through December 31, 2019) monthly incident rate of RSV infection. The model used a linear term to model time trend and Fourier terms to account for seasonality^14^. Using the model, we predicted the monthly incident rate of RSV infection during the pandemic months (January 1, 2020 through January 31, 2023). The predicted monthly incidence rate was compared with the observed rate during the same period by calculating rate ratios and 95% CIs using R, version 4.2.2. Data and code for time series analyses are available at https://github.com/liwang0904/RSV

#### Comparison with the CDC reported weekly rates of RSV-associated hospitalizations

We compared the EHR-based seasonality and time trend of monthly incidence rates of medically attended RSV infection in children aged 0**–**5 years of age with the CDC’s reported weekly rates of RSV-associated hospitalization among children aged 0**–**4 years of age from October 2018 through January 31, 2023. The CDC Respiratory Syncytial Virus Hospitalization Surveillance Network (RSV-NET) is a network that conducts active, population-based surveillance for laboratory-confirmed RSV-associated hospitalizations in children younger than 18 years of age and adults in 12 states covering almost 8% of the US^15^. Hospitalization rates are calculated as the number of residents in a surveillance area who are hospitalized with laboratory-confirmed RSV divided by the total population estimate for that area. State health departments are not required to report RSV-associated hospitalizations to the CDC. RSV-NET surveillance began tracking RSV-associated hospitalizations in children in the 2018–2019 season. Data were downloaded from the RSV-NET Surveillance System for children aged 0**–**4 years of age^16^.

#### Stratified analysis of seasonality

In this study, the medically encountered RSV infection was defined by 12 lab-test codes and 3 disease clinical diagnostic codes for RSV-associated diseases. The clinical diagnosis-based RSV infection included “Respiratory syncytial virus pneumonia” (J12.1), “Acute bronchiolitis due to respiratory syncytial virus” (J21.0), and “Respiratory syncytial virus as the cause of diseases classified elsewhere” (B97.4). We performed separate analysis for: clinical diagnosis-based RSV diseases (J12.1, J21.0, B97.4), positive lab test confirmed RSV infection (12 lab test codes) and RSV-associated bronchiolitis (J21.0). In addition, we also examined unspecified bronchiolitis (J21.9) to investigate whether the seasonality disruption was unique to RSV infection.

### 3. Statistical analysis in retrospective cohort study Study population

We examined whether prior COVID-19 infection was associated with increased risk of medically attended RSV infection among young children who had no prior RSV infection. For examining the association of prior COVID-19 infection with RSV infection in 2022 RSV season (10/2022– 12/2022) among children aged 0–5 years, the study population comprised 228,940 children of 0– 5 years (as of 10/2022) who had medical encounters with healthcare organizations in 10/2022 and had no prior medically attended RSV infection. The study population was then divided into two cohorts: (1) COVID-19 (+) cohort – 14,493 children who contracted COVID-19 any time prior to 8/2022 as documented in their EHR, and (2) COVID-19 (-) cohort – 214,447 children who had no documented COVID-19.

For examining the association of prior COVID-19 infection with first-time RSV infection in 2022 RSV season (10/2022–12/2022) among children aged 0–1 year, the study population comprised 99,105 children of 0–1 year (as of 10/2022) who had medical encounters with healthcare organizations in 10/2022 and had no prior medically attended RSV infection. The study population was then divided into two cohorts: (1) COVID-19 (+) cohort – 5,193 children who contracted COVID-19 prior to 8/2022 as documented in their EHR, and (2) COVID-19 (–) cohort – 93,912 children who had no documented COVID-19.

For examining the association of prior COVID-19 infection with first-time RSV infection in 2021 RSV season (7/2021–12/2021) among children aged 0–5 years, the study population comprised 370,919 children of 0–5 years (as of 7/2021–8/2021) who had medical encounters with healthcare organizations in 7/2021–8/2021and had no prior medically attended RSV infection. The study population was then divided into two cohorts: (1) COVID-19 (+) cohort – 6,309 children who contracted COVID-19 prior to 6/2022 as documented in their EHR, and (2) COVID-19 (–) cohort – 364,610 children who had no documented COVID-19.

For examining the association of prior COVID-19 infection with first-time RSV infection in 2021 RSV season (7/2021–12/2021) among children aged 0–1 year, the study population comprised 162,771 children of 0–1 year (as of 7/2021–8/2021) who had medical encounters with healthcare organizations in 7/2021–8/2021and had no prior medically attended RSV infection. The study population was then divided into two cohorts: (1) COVID-19 (+) cohort – 2,671 children who contracted COVID-19 prior to 6/2022 as documented in their EHR, and (2) COVID-19 (–) cohort – 160,100 children who had no documented COVID-19.

#### Statistical analysis

We examined the association of prior COVID-19 infection with first-time medically attended RSV infection occurring in 2022 RSV peak season (10/2022–12/2022) in young children by comparing the COVID-19 (+) and COVID-19 (−) cohorts. Cohorts were propensity-score matched (1:1 using a nearest neighbor greedy matching with a caliper of 0.25 times the standard deviation) for variables that are risk factors for RSV infection^17^ and are also potential risk factors for COVID-19^18^. The index event was a medical visit in 10/2022. Five outcomes were examined, including first-time medically attended overall RSV infection, positive lab test-confirmed RSV infection, clinically diagnosed RSV diseases, RSV-associated bronchiolitis, and unspecified bronchiolitis. The outcomes were followed for 60 days starting from the index event (a medical visit in 10/2022) and occurred from 10/1/2022–12/31/2022. Risk ratios (RR) and 95% confidence intervals were used to describe the relative risk of the outcomes between the matched COVID-19 (+) and COVID-19 (-) cohorts. Separate analyses were performed for children aged 0–5 years and children 0–1 year.

Similar cohort study was performed to examine the association of prior COVID-19 infection with first-time medically attended RSV infections occurring in 2021 RSV peak season (7/2021-12/2021) in young children by comparing the COVID-19 (+) and COVID-19 (−) cohorts. Cohorts were propensity-score matched (1:1) for variables that are risk factors for RSV infection^17^ and are also potential risk factors for COVID-19^18^. The index event was a medical visit in 7/2021–8/2021. Five outcomes were examined including first-time medically attended overall RSV infection, positive lab-test confirmed RSV, clinical diagnosis code-based RSV diseases, RSV-associated bronchiolitis, and unspecified bronchiolitis. The outcomes were followed for 120 days starting from the index event (medical visit in 7/2021–8/2021), which were outcomes that occurred in 7/1/2021–12/31/2021. Risk Ratios (RR) and 95% confidence intervals were used to calculate the relative risk of the outcomes. Separate analyses were performed for children aged 0–5 years and children 0–1 year.

Cohort study design and associated statistical tests were conducted within the TriNetX Advanced Analytics Platform. The TriNetX platform calculates RRs and associated CIs using R version 4.0.2. Cohort studies were conducted during 2/20/2023–3/5/2023 within the TriNetX Analytics Platform.

#### Data Availability Statement

This study used population-level aggregate and de-identified data generated by the TriNetX Platform. Due to data privacy, patient-level data were not used and cannot be shared.

#### Code Availability Statement

Data and code (R statistical software version 4.2.2) for time-series analysis are available at https://github.com/liwang0904/RSV. R 4.4.4 was used for calculating rate ratios of incidence rates. Cohort study design and associated statistics including propensity-score matching and measure of associations were conducted within the TriNetX Advanced Analytics Platform using built-in functions (R version 4.0.2) with significance set at p-value < 0.05 (two-sided). All R packages are freely available.

## References

1. Hall, C. B. et al. The burden of respiratory syncytial virus infection in young children. N. Engl. J. Med. 360, 588–598 (2009).

2. Olsen, S. J. et al. Changes in Influenza and Other Respiratory Virus Activity During the COVID-19 Pandemic - United States, 2020-2021. MMWR Morb. Mortal. Wkly. Rep. 70, 1013–1019 (2021).

3. Abbasi, J. “This Is Our COVID”—What Physicians Need to Know About the Pediatric RSV Surge. JAMA (2022) doi:10.1001/jama.2022.21638.

4. Willyard, C. Flu and colds are back with a vengeance — why now? Nature Publishing Group UK http://dx.doi.org/10.1038/d41586-022-03666-9 x(2022) doi:10.1038/d41586-022-03666-9.

5. Adams, G. et al. Viral Lineages in the 2022 RSV Surge in the United States. N. Engl. J. Med. (2023) doi:10.1056/NEJMc2216153.

6. Messacar, K. et al. Preparing for uncertainty: endemic paediatric viral illnesses after COVID-19 pandemic disruption. Lancet 400, 1663–1665 (2022).

7. Cohen, R. et al. Pediatric Infectious Disease Group (GPIP) position paper on the immune debt of the COVID-19 pandemic in childhood, how can we fill the immunity gap? Infect Dis Now 51, 418–423 (2021).

8. Billard, M.-N. & Bont, L. J. Quantifying the RSV immunity debt following COVID-19: a public health matter. The Lancet infectious diseases vol. 23 3–5 (2023).

9. Baker, R. E. et al. The impact of COVID-19 nonpharmaceutical interventions on the future dynamics of endemic infections. Proc. Natl. Acad. Sci. U. S. A. 117, 30547–30553 (2020).

10. Bardsley, M. et al. Epidemiology of respiratory syncytial virus in children younger than 5 years in England during the COVID-19 pandemic, measured by laboratory, clinical, and syndromic surveillance: a retrospective observational study. Lancet Infect. Dis. 23, 56–66 (2023).

11. Davis, H. E., McCorkell, L., Vogel, J. M. & Topol, E. J. Long COVID: major findings, mechanisms and recommendations. Nat. Rev. Microbiol. (2023) doi:10.1038/s41579-022-00846-2.

12. Iwasaki, A. & Putrino, D. Why we need a deeper understanding of the pathophysiology of long COVID. Lancet Infect. Dis. (2023) doi:10.1016/S1473-3099(23)00053-1.

13. Phetsouphanh, C. et al. Immunological dysfunction persists for 8 months following initial mild-to-moderate SARS-CoV-2 infection. Nat. Immunol. 23, 210–216 (2022).

14. Liu, J. et al. Analysis of the long-term impact on cellular immunity in COVID-19-recovered individuals reveals a profound NKT cell impairment. MBio 12, (2021).

15. Haunhorst, S. et al. A scoping review of regulatory T cell dynamics in convalescent COVID-19 patients – indications for their potential involvement in the development of Long COVID? Front. Immunol. 13, (2022).

16. RSV-NET: Respiratory Syncytial Virus Hospitalization Surveillance Network, Centers for Disease Control and Prevention. https://www.cdc.gov/rsv/research/rsv-net/dashboard.html (2023).

17. CDC. Centers for Disease Control and Prevention. Wearing Masks in Travel and Public Transportation Settings. Centers for Disease Control and Prevention https://www.cdc.gov/coronavirus/2019-ncov/travelers/masks-public-transportation.html (2022).

18. Children and COVID-19: State-Level Data Report. https://www.aap.org/en/pages/2019-novel-coronavirus-covid-19-infections/children-and-covid-19-state-level-data-report/ (2022).

19. Wang, L. et al. Incidence Rates and Clinical Outcomes of SARS-CoV-2 Infection With the Omicron and Delta Variants in Children Younger Than 5 Years in the US. JAMA Pediatr. (2022) doi:10.1001/jamapediatrics.2022.0945.

20. Justice, N. A. & Le, J. K. Bronchiolitis. (StatPearls Publishing, 2022).

21. Beňová, K., Hancková, M., Koči, K., Kúdelová, M. & Betáková, T. T cells and their function in the immune response to viruses. Acta Virol. 64, 131–143 (2020).

22. Rogers, M. C. et al. CD4+ regulatory T cells exert differential functions during early and late stages of the immune response to respiratory viruses. J. Immunol. 201, 1253–1266 (2018).

23. Piret, J. & Boivin, G. Viral Interference between Respiratory Viruses. Emerg. Infect. Dis. 28, 273–281 (2022).

24. Eden, J.-S. et al. Off-season RSV epidemics in Australia after easing of COVID-19 restrictions. Nat. Commun. 13, 2884 (2022).

25. Groves, H. E. et al. The impact of the COVID-19 pandemic on influenza, respiratory syncytial virus, and other seasonal respiratory virus circulation in Canada: A population-based study. The Lancet Regional Health - Americas 1, 100015 (2021).

26. Sommer, C., Resch, B. & Simões, E. A. F. Risk factors for severe respiratory syncytial virus lower respiratory tract infection. Open Microbiol. J. 5, 144–154 (2011).

27. Paul, L. A. et al. Association of Age and Pediatric Household Transmission of SARS-CoV-2 Infection. JAMA Pediatr. 175, 1151–1158 (2021).

## References

1. TriNetX. TriNetX https://trinetx.com/.

2. Wang, L. et al. Incidence Rates and Clinical Outcomes of SARS-CoV-2 Infection With the Omicron and Delta Variants in Children Younger Than 5 Years in the US. JAMA Pediatr. (2022) doi:10.1001/jamapediatrics.2022.0945.

3. Wang, L., Davis, P. B., Kaelber, D. C., Volkow, N. D. & Xu, R. Comparison of mRNA-1273 and BNT162b2 Vaccines on Breakthrough SARS-CoV-2 Infections, Hospitalizations, and Death During the Delta-Predominant Period. JAMA (2022) doi:10.1001/jama.2022.0210.

4. Wang, L., Davis, P. B., Kaelber, D. C. & Xu, R. COVID-19 breakthrough infections and hospitalizations among vaccinated patients with dementia in the United States between December 2020 and August 2021. Alzheimers. Dement. (2022) doi:10.1002/alz.12669.

5. Wang, W., Kaelber, D. C., Xu, R. & Berger, N. A. Breakthrough SARS-CoV-2 Infections, Hospitalizations, and Mortality in Vaccinated Patients With Cancer in the US Between December 2020 and November 2021. JAMA Oncol (2022) doi:10.1001/jamaoncol.2022.1096.

6. Wang, L., Kaelber, D. C., Xu, R. & Berger, N. A. COVID-19 breakthrough infections, hospitalizations and mortality in fully vaccinated patients with hematologic malignancies: A clarion call for maintaining mitigation and ramping-up research. Blood Rev. 100931 (2022).

7. Wang, L., Berger, N. A. & Xu, R. Risks of SARS-CoV-2 Breakthrough Infection and Hospitalization in Fully Vaccinated Patients With Multiple Myeloma. JAMA Netw Open 4, e2137575 (2021).

8. Wang, L. et al. Association of COVID-19 with new-onset Alzheimer’s disease. J. Alzheimers. Dis. (2022) doi:10.3233/JAD-220717.

9. Pan, Y., Davis, P. B., Kaebler, D. C., Blankfield, R. P. & Xu, R. Cardiovascular risk of gabapentin and pregabalin in patients with diabetic neuropathy. Cardiovasc. Diabetol. 21, 170 (2022).

10. Kendall, E. K., Olaker, V. R., Kaelber, D. C., Xu, R. & Davis, P. B. Association of SARS-CoV-2 Infection With New-Onset Type 1 Diabetes Among Pediatric Patients From 2020 to 2021. JAMA Netw Open 5, e2233014 (2022).

11. Wang, L., Xu, R., Kaelber, D. C. & Berger, N. A. Time Trend and Association of Early-Onset Colorectal Cancer with Diverticular Disease in the United States: 2010-2021. Cancers 14, (2022).

12. Wang L, Wang Q, Davis PB, Volkow ND, Xu R. Increased risk for COVID-19 breakthrough infection in fully vaccinated patients with substance use disorders in the United States between December 2020 and August 2021. World Psychiatry 21, 124–132 (2022).

13. Wang, L. et al. Association of COVID-19 with endocarditis in patients with cocaine or opioid use disorders in the US. Mol. Psychiatry (2022) doi:10.1038/s41380-022-01903-1.

14. Ord, K., Fildes, R. A. & Kourentzes, N. Principles of business forecasting. https://eprints.lancs.ac.uk/id/eprint/88208/1/POBF_2e_handout_pages.pdf (2017).

15. RSV-NET: Respiratory Syncytial Virus Hospitalization Surveillance Network, Centers for Disease Control and Prevention. https://www.cdc.gov/rsv/research/rsv-net/dashboard.html (2023).

16. Weekly Rates of Laboratory-Confirmed RSV Hospitalizations from the RSV-NET Surveillance System. (2022).

17. Sommer, C., Resch, B. & Simões, E. A. F. Risk factors for severe respiratory syncytial virus lower respiratory tract infection. Open Microbiol. J. 5, 144–154 (2011).

18. CDC. Underlying Medical Conditions Associated with Higher Risk for Severe COVID-19: Information for Healthcare Professionals. Centers for Disease Control and Prevention https://www.cdc.gov/coronavirus/2019-ncov/hcp/clinical-care/underlyingconditions.html (2023).

